# Clinical Progression on CDR-SB ^©^: Progression Free Time at Each 0.5-unit Level in Dominantly Inherited and Sporadic Alzheimer’s Disease Populations

**DOI:** 10.1101/2025.02.17.25322322

**Authors:** Guoqiao Wang, Yan Li, Eric McDade, Chengjie Xiong, Sarah M. Hartz, Randall J. Bateman, John C. Morris, Lon S. Schneider

**Affiliations:** Department of Neurology, Washington University, School of Medicine, St. Louis, 63130, United States; Division of Biostatistics, Washington University School of Medicine in St. Louis, 660 S. Euclid Ave, St. Louis, MO 63110; Department of Psychiatry, Washington University School of Medicine in St. Louis, 660 S. Euclid Ave, St. Louis, MO 63110; Department of Psychiatry and The Behavioral Sciences, Department of Neurology, Keck School of Medicine, University of Southern California, Los Angeles, 90033, United States

**Keywords:** Progression-free time, Alzheimer disease, CDR-SB, clinically meaningful interpretation, disease progression model

## Abstract

**INTRODUCTION:** CDR-SB is a reliable and clinically meaningful composite for assessing treatment effects in Alzheimer’s disease (AD) clinical trials. Small CDR-SB differences at the end of a trial often lead to controversy in deriving clinically meaningful interpretations.

**METHODS:** We estimated progression-free time participants remained at each 0.5-unit CDR-SB increment in dominantly inherited AD (DIAD) and sporadic AD populations, evaluating its potential as an alternative measure of treatment effects.

**RESULTS:** Progression-free time is longer at CDR-SB ≤ 2.0 (1–2 years) and shorter at CDR-SB ≥ 5 (0.33 or less) in the ADNI cohort. The DIAD cohort showed similar but shorter times. Using progression-free time, continuous lecanemab treatment for three years is estimated to delay disease progression by 0.7 years in the sporadic population.

**DISCUSSION:** Progression-free time provides a benchmark for expressing clinical progression and treatment effects and can be applied particularly during open-label extensions and singlearm trials without placebo comparisons.

## 1 Background

The Clinical Dementia Rating Scale - Sum of Boxes (CDR-SB^©^) is widely used as a primary or secondary endpoint in Phase II and III clinical trials for Alzheimer’s disease (AD).^1-5^ It serves as an FDA-approved composite measure for assessing mild cognitive impairment (MCI) and mild dementia, simplifying trials that might otherwise require multiple or co-primary endpoints.^6^

In the CDR-SB assessment, a rater uses semi-structured interviews of the participant and an informant to assess the former’s performance across six domains: three cognitive (memory, orientation, judgment/problem-solving) and three functional (community affairs, home/hobbies, and personal care). Each domain is scored on an ordinal scale (0, 0.5, 1, 2, 3), indicating ‘no dementia,’ ‘uncertain,’ ‘mild,’ ‘moderate,’ and ‘severe’ dementia respectively, with the exception of personal care, which omits the 0.5 score. The scores across these domains are summed to yield the CDR-SB score which has a range of 0 to 18 with 0.5-point intervals at lower summary scores (i.e., better function).

CDR-SB scores and the differences in change from baseline between treatment and placebo groups are prominently presented in the clinical studies sections of the prescribing information (i.e., “the label”) for the FDA approved amyloid-targeting antibodies, aducanumab,^1^ lecanemab,^3^ and donanemab,^4^ providing patients and prescribers with insights into treatment outcomes. Although treatment-placebo differences of the CDR-SB are statistically significant for these antibodies and supported FDA’s approval of these antibodies for marketing, the actual mean effect sizes are modest, ranging from +0.03,^1^ to -0.39,^1^ -0.45,^3^ and -0.70,^4^ sparking considerable debate regarding their clinical interpretability and significance. Understanding how these numerical differences might translate into meaningful clinical outcomes is essential for clinicians, patients, and researchers alike. An Alzheimer’s Association workgroup suggested that expectations of outcomes from therapeutic interventions in AD may need to be modified.^7^

One approach is to express differences in CDR-SB changes as a percentage reduction compared to the placebo comparator, e.g. x% slowing of progression.^2-4^ Another is to interpret these differences as a delay in disease progression, either relative to the decline observed in the placebo group (e.g., 5.3 months for lecanemab)^4,8^ or relative to reaching a specific disease stage milestone.^9^ However, these methods are *relative* measures, vary with the change in placebo, and may not be applicable during the open-label extension (OLE) period when all participants receive treatment. Furthermore, these approaches are trial-specific and may lack comparability across trials due to differences in baseline CDR-SB levels and change in the placebo control groups.

In this study, we introduce an alternative methodology that standardizes the evaluation of delays in disease progression based on the CDR scale. We accomplish this by calculating the progression-free time (or ‘residence time’) a research participant remains at a given CDR-SB level before progressing to a higher level. Specifically, we propose estimating the progression-free time at each 0.5-unit CDR-SB increment for two distinct populations: participants with dominantly inherited Alzheimer’s disease (DIAD) from the Dominantly Inherited Alzheimer Network (DIAN) observational study and those with sporadic Alzheimer’s disease from the Alzheimer’s Disease Neuroimaging Initiative (ADNI) study. We suggest using progression-free time, or time-to-0.5-unit progression, as a potential measure of clinically meaningful change.

## 2 Methods

### 2.1 Study Participants

The Dominantly Inherited Alzheimer Network (DIAN) is an international, multicenter observational study focused on individuals aged 18 years or older who have parents with a known causative mutation in the *amyloid precursor protein (APP), presenilin 1 (PSEN1)*, or *presenilin 2 (PSEN2)* genes, for AD. Mutation carriers (MCs) face an almost certain risk of developing AD, with relatively predictable symptom onset ages. At enrollment into the DIAN cohort, these participants were either cognitively unimpaired (Clinical Dementia Rating [CDR] = 0) or showed signs of early symptomatic disease (CDR = 0.5 or 1, signifying very mild or mild dementia, respectively). Research participants with the Dutch mutation were excluded in this study due to their atypical rates of clinical progression. Detailed information on the DIAN study’s structure and the assessments performed can be found in previous publications.^10,11^

The Alzheimer’s Disease Neuroimaging Initiative (ADNI) is a longitudinal, multicenter study designed to track the progression of Alzheimer’s disease using various biomarkers in individuals aged 55 to 90 years. ADNI encompasses participants across the spectrum of AD pathology, categorizing them into three groups at enrollment into ADNI: normal cognition, mild cognitive impairment (MCI), and AD dementia. Data included in this study were download from adni.loni.ucla.edu.

### 2.2 Statistical Analysis

To map the holistic trajectory of disease progression from CDR-SB score of 0 (the lower bound) to a CDR-SB score of 18 (the upper bound), we applied a parametric disease progression model incorporating random effects for both the endpoint and the time variable (see Supplemental Material for details). In alignment with previous studies,^12-14^ integrating a random effect into the time variable enables the model to accurately recover the underlying disease progression pathway by adjusting the time variable: shifting it leftward for less severe stages and rightward for more severe stages of the disease. For mutation carriers in the DIAN study, the Estimated Years from Symptom Onset (EYO) serves as a relatively precise indicator of their disease stage, thus, EYO was adopted as the time variable in our model.^12^ Conversely, for participants in the ADNI study, lacking such an accurate measure, the time since enrollment in years was used as the time variable.^13,14^ Following the estimation of the disease progression trajectory, the progression-free time—the duration spent at each 0.5-unit interval of CDR-SB progression— was calculated based on the rate of disease progression observed in each interval.

To demonstrate the potential use of progression-free time, we assessed the treatment effect of lecanemab during the OLE period as a time delay in disease progression.^15^ This was followed by a comparison with the delay in disease progression as estimated by conventional methods, such as calculating the time based on the percentage reduction in disease progression compared to placebo, multiplied by the total duration.^8^

All statistical analyses were conducted using SAS software, version 9.4. We utilized two-sided t-tests with a Type I error rate of 5% and reported the corresponding 95% confidence intervals (CIs).

## 3 Results

### 3.1 Progression-free Time for DIAD and Sporadic AD Populations

In the DIAN study, 384 mutation carriers were included from data freeze 17 (without Dutch mutations), while in the ADNI study, 406 participants were involved. The baseline characteristics of these participants are detailed in **Table 1**.

**Table 1:** Baseline characteristics for each study.

| Characteristic | ADNI (N=406) | DIAN (N=384) |
| --- | --- | --- |
| CDR = 0, N (%) | 210 (51.7%) | 242 (63.0%) |
| CDR > 0, N (%) | 196 (48.3%) | 142 (37.0%) |
| Age, years (SD) | 72.9 ± 7.3 | 38.2 ± 7.3 |
| MMSE | 28.5 ± 1.9 | 26.5 ± 5.5 |
| APOE 4 Negative, N (%) | 267 (65.8%) | 271 (70.6%) |
| CDR-SB, (SD) | 0.6 ± 1.0 | 1.5 ± 3.1 |
| Follow-up duration, years (SD) | 5.4 (1.8) | 4.1 (2.6) |

Figure 1 illustrates the progression-free time at each 0.5-unit increment of the CDR-SB score, up to 12.5, for the DIAD population. The progression-free time at CDR-SB 0.5 is approximately 1.65 years, which sharply declines to less than 0.5 years once the CDR-SB score reaches 2.5 or above Supplemental Table 1 presents the progression-free time for all intervals at 0.5-unit increments of the CDR-SB score.

**Figure 1:**
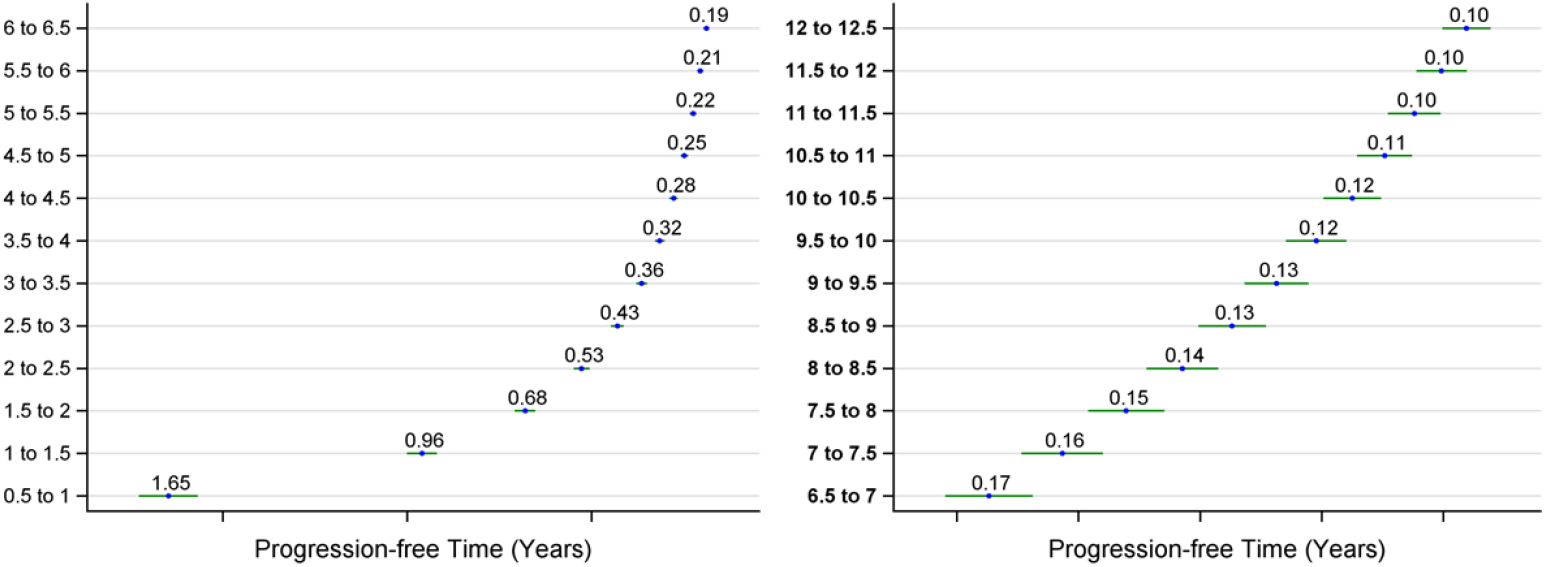
Estimated progression-free time and its 95% CI for each 0.5-unit CDR-SB interval based on the DIAN study.

Figure 2 presents similar data for the sporadic AD population, showing progression-free times for each 0.5-unit increment of the CDR-SB score, also up to 12.5. Here, the progression-free time from CDR-SB 0.5 to CDR-SB 1 is roughly 2.25 years, which then rapidly decreases to under 0.5 years when the CDR-SB score surpasses 3.5. The progression-free time for all intervals at 0.5-unit increments of the CDR-SB score are presented in Supplemental Table 1.

**Figure 2:**
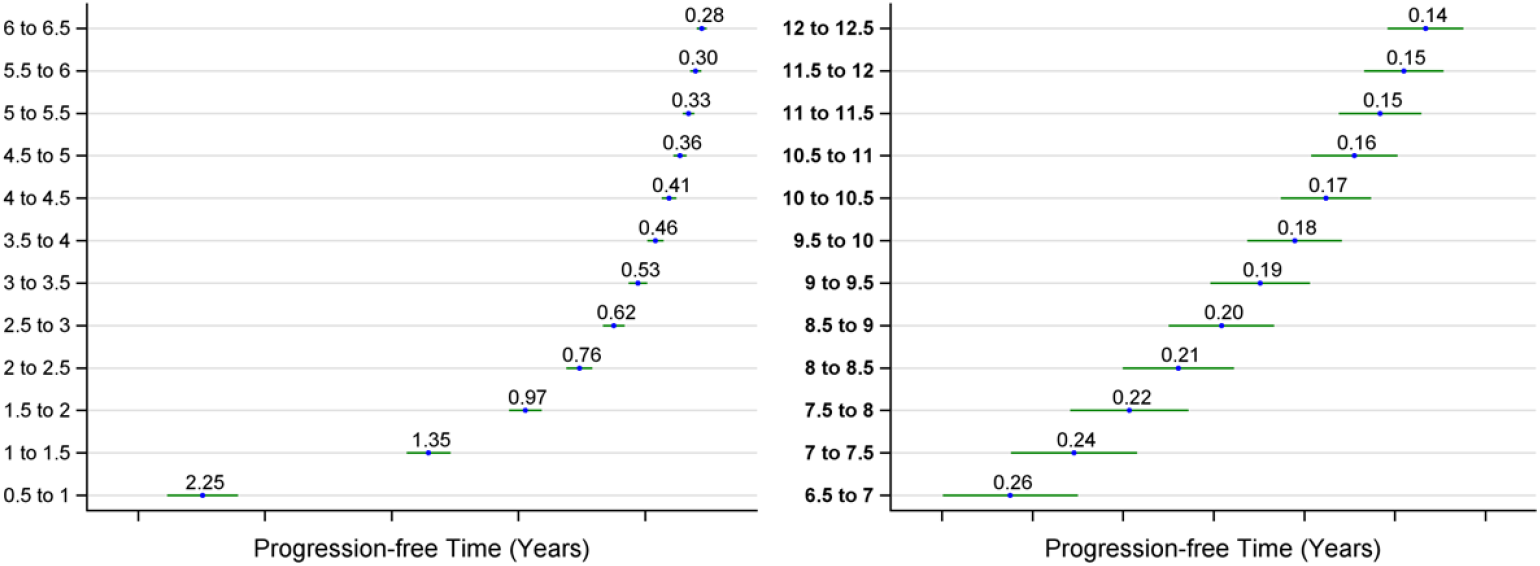
Estimated progression-free time and its 95% CI for each 0.5-unit CDR-SB interval based on the ADNI study.

### 3.2 Application of Progression-free Time to Assess Treatment Effects

According to reports from the OLE period of the phase III lecanemab trial, participants in the early-start group, who began treatment from the double-blind baseline, showed a decline of 3.09 units in the CDR-SB score over 3 years, moving from a baseline of 3.17 to 6.26.^3,15^ Since participants in the original placebo group transitioned to lecanemab treatment during the OLE (becoming the delayed-start group), it’s not feasible to directly assess the treatment’s effect in the early-start group by comparing it with the delayed-start group during this period. Instead, we could evaluate the treatment effect using the estimated progression-free time (Figure 2) for the sporadic AD population.

Our estimation suggests that without lecanemab treatment, participants might have progressed from a CDR-SB score of 3.17 to 6.26 in just 2.3 years. However, with continuous lecanemab treatment, this progression was extended to 3 years, effectively a time difference or delay of progression of 0.7 years.

As an alternative approach to assess the treatment effect, a cohort from the ADNI study, matched as external controls, was used to benchmark disease progression from the double-blind baseline to the 36-month mark, including 18 months within the OLE period.^15^ It was observed that the matched ADNI controls experienced a decline of 4.14 units in the CDR-SB score, which was 0.95 units greater than the decline in the early-start group receiving lecanemab.^15^ To quantify the delay in disease progression using the percentage reduction method,^5^ we first calculated the percentage reduction in disease progression relative to the ADNI group. We then multiplied this value by the total study duration in years. This calculation yielded a delay of 0.7 years, derived from the formula (0.95/4.14) x (36/12),^5^ which aligns well with the findings obtained using the progression-free time approach.

## 4 Discussion

CDR-SB has been demonstrated as a reliable composite measure for assessing treatment effects in Alzheimer’s disease clinical trials and its use in AD drug development is ongoing.^1,3,4^ Consequently, there is a great interest among clinicians, patients, and researchers to derive meaningful interpretations from the treatment effects measured by the change over time and treatment-control differences in CDR-SB in trials.^7^ Various approaches have been suggested to quantify these effects, including: (i) Expressing the difference in CDR-SB scores between treatment and control groups as mean differences; (ii) in terms of percentage reduction compared to controls ;^3,4^ (iii) translating these differences into time savings relative to the decline observed in the control or placebo group;^4,8,16^ (iv) calibrating the treatment effect to reflect delays in reaching critical milestones, such as the loss of independence in activities of daily living.^9^ These methodologies have significantly enhanced our ability to interpret the outcomes of recent amyloid-targeting antibody trials.^3,4^ Building on these, our study introduces an alternative approach by calculating the progression-free time for each 0.5-unit increment in the CDR-SB score for both DIAD and sporadic AD populations.

Our findings indicate that the DIAD population progresses faster than the sporadic AD population, consistent with previous reports based on cognitive endpoints.^17^ Applying this progression-free-time approach to quantify treatment effects in the Clarity AD trial during its 18-month OLE period yielded an estimated 0.7-year delay in disease progression, aligning closely with results from the existing method proposed by Wang. et al.^5^ This suggests that progression-free time, or ‘residence time,’ at a given 0.5-unit increment in CDR-SB can provide a standardized measure of treatment effects.

The concept also offers clinical relevance: during early AD, patients with lower CDR-SB scores (e.g., 0.5–1.0) may remain stable over 1–2 years, making even a 0.5-point decrease clinically significant. Conversely, at higher CDR-SB scores (e.g., ≥2.5), progression-free time shortens to approximately 0.5–0.6 years, where a 1.0-point reduction may represent a minimal clinically important difference. By quantifying the time patients remain at specific disease stages, progression-free time provides a useful tool for clinical interpretation of treatment effects over extended periods, particularly during OLE periods, and in single-arm trials where placebo comparisons are unavailable.

Our study represents an initial attempt to quantify progression-free time for each CDR-SB 0.5 unit; however, several limitations should be considered. First, our estimates are based on a parametric model, which, although fitting the data reasonably well, requires further validation to assess its robustness. Future studies should explore alternative models such as time-to-event based models to confirm the consistency of our findings. Second, our results need validation using different study cohorts to assess variability. Finally, we did not report progression-free time from CDR-SB 0 to CDR-SB 0.5 because an individual’s CDR-SB 0 stage can last for many years, and the estimated transition time depends on when participants enter the study. Future research could explore progression-free time from an earlier benchmark, such as amyloid positivity in cognitively normal individuals, to CDR-SB 0.5, providing a clearer timeline of disease progression during this stage.

Much like oncology’s use of progression-free survival, a progression-free time chart could serve as a benchmark for understanding AD progression. This approach would allow clinicians to communicate the rate of cognitive decline in a more accessible and intuitive manner, facilitating clearer discussions with patients and caregivers. By framing disease progression in terms of the time delayed in disease progression by stage of illness, or the potential extension of stability offered by treatment, this method translates abstract concepts into tangible metrics that may be more interpretable by patients, families and physicians. For patients, families, and clinicians such a chart could simplify the understanding of disease progression and treatment impacts, enhancing engagement and adherence to treatment protocols. More informed discussions about AD treatment effects may be provided by integrating progression-free time metrics into clinical practice, helping set realistic expectations and plan for future care needs.

## Data Availability

All data produced are available online at:
https://adni.loni.usc.edu/data-samples/adni-data/
https://dian.wustl.edu/observational-study/

## Supplemental Materials

### Parametric disease progression model (DPM)

Briefly, the model applied to the DIAN study can be described as:

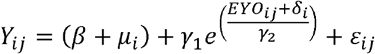

Where,

- *Y*_*ij*_ denotes the assessment for subject *i* at visit *j* (*EYO*_*ij*_)
- *β* is the mean of the healthy period (e.g. EYO <=-15)
- *γ*_1_ is the shape parameter and influences how much steep the curve exhibits. It also represents the mean decline from the healthy period to symptom onset (e.g. EYO=0)
- *γ*_2_ represents the scale parameter, and affects the rate at which the exponential function grows or decays
- *μ*_*i*_ is the subject-level random effect that shifts the individual data up or down on the y-axis, accounting for individual variability in the baseline or intercept of the response variable
- *δ*_*i*_ is the subject-level random effect that shifts the individual data left or right on the x-axis, adjusting for individual variability in the timing or progression from symptom onset
- *μ*_*i*_ and *δ*_*i*_ are assumed to follow a bivariate normal distribution: 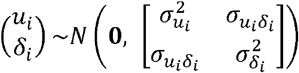
- *ε*_*ij*_ is the within-subject error and is assumed to follow a normal distribution 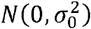 for participants with CDR 0 at baseline and 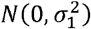 for participants with CDR>0.

For ADNI study, the same model is applied with EYO being replaced by the time since enrollment in years.

After the DPM is estimated, the progression time to each CDR-SB level was calculated based on the model. Then the progression free time was calculated as the difference between the progression time corresponding to two consecutive CDR-SB levels.

**Supplemental Table 1:** Estimated progression-free time for each 0.5-unit CDR-SB interval for each study.

|  | DIAN Study |  |  |  | ADNI Study |  |  |  |
| --- | --- | --- | --- | --- | --- | --- | --- | --- |
| CDR-SB Interval | Estimate | SE | Lower | Upper | Estimate | SE | Lower | Upper |
| 0.5 to 1 | 1.647 | 0.041 | 1.567 | 1.728 | 2.250 | 0.071 | 2.106 | 2.387 |
| 1 to 1.5 | 0.960 | 0.021 | 0.919 | 1.001 | 1.350 | 0.044 | 1.268 | 1.441 |
| 1.5 to 2 | 0.680 | 0.014 | 0.652 | 0.708 | 0.970 | 0.033 | 0.908 | 1.037 |
| 2 to 2.5 | 0.527 | 0.011 | 0.506 | 0.549 | 0.760 | 0.026 | 0.708 | 0.811 |
| 2.5 to 3 | 0.431 | 0.009 | 0.413 | 0.448 | 0.620 | 0.022 | 0.580 | 0.666 |
| 3 to 3.5 | 0.364 | 0.008 | 0.349 | 0.379 | 0.530 | 0.019 | 0.492 | 0.565 |
| 3.5 to 4 | 0.315 | 0.007 | 0.302 | 0.328 | 0.460 | 0.016 | 0.426 | 0.491 |
| 4 to 4.5 | 0.278 | 0.006 | 0.267 | 0.289 | 0.410 | 0.015 | 0.376 | 0.434 |
| 4.5 to 5 | 0.249 | 0.005 | 0.238 | 0.259 | 0.360 | 0.013 | 0.337 | 0.389 |
| 5 to 5.5 | 0.225 | 0.005 | 0.216 | 0.234 | 0.330 | 0.012 | 0.305 | 0.352 |
| **5.5 to 6** | 0.205 | 0.004 | 0.197 | 0.214 | 0.300 | 0.011 | 0.279 | 0.322 |
| **6 to 6.5** | 0.189 | 0.004 | 0.181 | 0.197 | 0.280 | 0.010 | 0.256 | 0.297 |
| **6.5 to 7** | 0.175 | 0.004 | 0.168 | 0.182 | 0.260 | 0.009 | 0.238 | 0.275 |
| **7 to 7.5** | 0.163 | 0.003 | 0.156 | 0.169 | 0.240 | 0.009 | 0.221 | 0.256 |
| **7.5 to 8** | 0.152 | 0.003 | 0.146 | 0.158 | 0.220 | 0.008 | 0.207 | 0.240 |
| **8 to 8.5** | 0.143 | 0.003 | 0.137 | 0.149 | 0.210 | 0.008 | 0.194 | 0.225 |
| **8.5 to 9** | 0.135 | 0.003 | 0.129 | 0.140 | 0.200 | 0.007 | 0.183 | 0.212 |
| **9 to 9.5** | 0.128 | 0.003 | 0.122 | 0.133 | 0.190 | 0.007 | 0.174 | 0.201 |
| **9.5 to 10** | 0.121 | 0.003 | 0.116 | 0.126 | 0.180 | 0.007 | 0.165 | 0.191 |
| **10 to 10.5** | 0.115 | 0.002 | 0.110 | 0.120 | 0.170 | 0.006 | 0.157 | 0.182 |
| **10.5 to 11** | 0.110 | 0.002 | 0.105 | 0.114 | 0.160 | 0.006 | 0.149 | 0.173 |
| **11 to 11.5** | 0.105 | 0.002 | 0.100 | 0.109 | 0.150 | 0.006 | 0.143 | 0.166 |
| **11.5 to 12** | 0.100 | 0.002 | 0.096 | 0.105 | 0.150 | 0.006 | 0.137 | 0.159 |
| **12 to 12.5** | 0.096 | 0.002 | 0.092 | 0.100 | 0.140 | 0.005 | 0.131 | 0.152 |
| **12.5 to 13** | 0.092 | 0.002 | 0.089 | 0.096 | 0.140 | 0.005 | 0.126 | 0.146 |
| **13 to 13.5** | 0.089 | 0.002 | 0.085 | 0.093 | 0.130 | 0.005 | 0.121 | 0.141 |
| **13.5 to 14** | 0.086 | 0.002 | 0.082 | 0.089 | 0.130 | 0.005 | 0.117 | 0.136 |
| **14 to 14.5** | 0.083 | 0.002 | 0.079 | 0.086 | 0.120 | 0.005 | 0.113 | 0.131 |
| **14.5 to 15** | 0.080 | 0.002 | 0.077 | 0.083 | 0.120 | 0.004 | 0.109 | 0.126 |
| **15 to 15.5** | 0.077 | 0.002 | 0.074 | 0.080 | 0.110 | 0.004 | 0.105 | 0.122 |
| **15.5 to 16** | 0.075 | 0.002 | 0.072 | 0.078 | 0.110 | 0.004 | 0.102 | 0.118 |
| **16 to 16.5** | 0.073 | 0.002 | 0.070 | 0.076 | 0.110 | 0.004 | 0.099 | 0.115 |
| **16.5 to 17** | 0.070 | 0.001 | 0.067 | 0.073 | 0.100 | 0.004 | 0.096 | 0.111 |
| **17 to 17.5** | 0.068 | 0.001 | 0.065 | 0.071 | 0.100 | 0.004 | 0.093 | 0.108 |
| **17.5 to 18** | 0.066 | 0.001 | 0.064 | 0.069 | 0.100 | 0.004 | 0.091 | 0.105 |

## DIAN Observational Study Site Investigators

James Noble, MD, Columbia University, New York

Martin Farlow, MD, Indiana University, Indianapolis, IN

Jasmeer Chhatwal, MD, PhD, Brigham and Women’s Hospital-Massachusetts GH, Charlestown, MA

Stephen Salloway, MD, Butler Hospital, Warren Alpert School of Medicine, Brown University

Sarah Berman, MD, PhD, University of Pittsburgh, PA

Gregg Day, MD, Mayo Clinic Jacksonville, FL

Hiroyuki Shimada, MD, PhD, Osaka City University, Japan

Takeshi Ikeuchi, MD, PhD, Brain Research Institute, Nigata, Japan

Kazushi Suzuki, MD, PhD, The University of Tokyo, Japan

Peter Schofield, PhD, DSc, Neuroscience Research Australia, Sydney

Ralph Martins, BSc, PhD, Edith Cowan University, Nedlands, Western Australia

Nick Fox, MD, FRCP, FMedSci, Dementia Research Centre, University College London, United Kingdom

Johannes Levin, MD, PhD, German Center for Neurodegenerative Diseases (DZNE), Munich, Germany

Mathias Jucker, PhD, German Center for Neurodegenerative Diseases (DZNE), Tubingen, Germany

Raquel Sanchez Valle, MD, Hospital Clinic i Provincial de Barcelona, Spain

Patricio Chrem, MD, Fundación para la Lucha contra las Enfermedades Neurológicas de la Infancia (FLENI), Buenos Aires, Argentina

## DIAN EXR Referring Clinicians, Researchers and Partner Sites

Neelum T. Aggarwal, MD, Rush University Medical Center, Chicago IL

Tom Ala, Center for Alzheimer’s Disease and Related Disorders, Southern Illinois University

School of Medicine

Thomas Bird, University of Washington, Seattle

Sandra E. Black, Sunnybrook Health Sciences Centre, University of Toronto, Canada

William J. Burke, MD, Banner Alzheimer’s Institute

Cynthia M. Carlsson, MD, MS, University of Wisconsin School of Medicine and Public Health

Andrew Frank M.D. B.Sc.H. F.R.C.P.(C), Bruyere Continuing Care, Ottawa, Ontario, Canada

James E. Galvin, MD, MPH, Charles E. Schmidt College of Medicine, Florida Atlantic University

Alvin C Holm, MD, Bethesda Hospital, St. Paul, MN

John S.K. Kauwe, Brigham Young University

David Knopman MD, Mayo Clinic, Rochester MN

Sarah Kremen, MD, University of California, Los Angeles

Alan J. Lerner, University Hospitals Cleveland Medical Center

Barry S. Oken, MD, PhD, Oregon Health & Science University

Hamid R. Okhravi, Eastern Virginia Medical School

Ronald C. Petersen, Mayo Clinic, Rochester, MN

Aimee L. Pierce, MD, University of California Irvine

Marsha J. Polk, MED, University of Texas Health Science Center at San Antonio

John M. Ringman, MD, MS, University Southern California

Peter St. George Hyslop, MD, FRS, FRSC, FRCPC, University of Toronto

Sanjeev N. Vaishnavi, MD, PhD, University of Pennsylvania

Sandra Weintraub, Northwestern University Feinberg School of Medicine, IL

